# Retrospective Case-Control Study of Pre-Diagnosis Observational and Prescription Data in Parkinson’s Disease

**DOI:** 10.1101/2025.08.11.25333439

**Authors:** Patrick Doherty, Susan Duty, Gareth Williams

## Abstract

**Background:** There are still no disease-modifying treatments available for Parkinson’s disease (PD) despite the growing interest in trialling repurposed drugs. Retrospective analysis of real-world health records may identify early predictive biomarkers and therapeutic candidates for increasing the chances of a positive trial outcome.

**Methods:** We conducted a retrospective case-control study using the UK-wide Clinical Practice Research Datalink (CPRD), comparing biometric and prescription data from over 38,000 individuals diagnosed with PD to 114,000 matched controls, across 5 to 10 and 10 to 20 years prior to diagnosis.

**Results:** The PD cohort was characterised by significant deviations in established biometric risk factors, including markedly elevated systolic arterial pressure, serum urea, and creatinine levels, alongside widespread alterations in metabolic and inflammatory markers. A combined risk score provided a moderate predictive accuracy (ROC AUC = 0.73) in line with published analyses. We confirmed the previously reported contrasting associations of salbutamol and propranolol with PD incidence. We further found hormonal drugs and triptan migraine medication associated with a lower PD incidence. However, these drugs had a higher prescription rate in those with lower biometric-based risk possibly confounding their positive effects against PD risk. In contrast GLP-1 receptor agonists tended to be prescribed in those with higher underlying PD risk but were associated with a lower PD incidence.

**Conclusion:** Integrating biometric and prescription histories enables more accurate interpretation of medication-disease associations, highlighting GLP-1 receptor agonists as strong candidates for PD intervention. Our findings support a data-driven framework for early risk detection and therapeutic discovery in neurodegenerative diseases.

## Introduction

Parkinson’s disease (PD) is the second most common neurological condition after Alzheimer’s disease (AD), with more than 10 million people currently living with PD, worldwide. PD manifests as a combination of motor and non-motor symptoms and is pathologically characterised by loss of dopaminergic neurones in the nigrostriatal tract and alpha synuclein pathology in the form of Lewy bodies and Lewy neurites which spread throughout the brain. The established nigrostriatal pathology of PD allows for effective management regimens through dopamine replacement compensating for the loss of dopaminergic neurons [1]. However, while helping to better control movement, this symptomatic approach does not address the progressive neuronal loss or many of the non-motor symptoms including the later development of dementia[2]. Like AD, PD is characterized by a complex pathogenesis involving a combination of neurodegeneration, protein misfolding [3], inflammation[4], and mitochondrial dysfunction[5]. Also like AD, multiple weak genetic associations with risk have been discovered[6], with evidence for a degree of commonality across neurodegenerative conditions[7]. The environmental[8] and lifestyle risk factors[9] associated with late onset are also common across neurodegenerative diseases. This begs the question whether PD should be classified as a disease at all, suggesting instead that, given the discrepancy between its underlying causes and clinical manifestations, it may be more accurately described as a syndrome[10], as has more recently been proposed for AD [11]. This multifactorial landscape of PD and dementia in general has prompted interest in alternative strategies for the discovery of novel therapeutics, particularly drug repurposing, where approved medications with known safety profiles are explored for efficacy in new indications[12, 13]. Alternative application of existing medications may result from the discovery of shared targets[12] or the commonalities emerging from the analysis of high content biological data related to drug activity and disease phenotype, such as gene expression profiles[14, 15]. Retrospective analysis of prescription histories and disease incidence can lead to the emergence of promising candidates, such as the case of salbutamol identified as a potential protective agent against PD through an analysis of the Norwegian NorPD prescription database [16]. While this association does not establish causality it offers a clear mechanistic hypothesis for future investigation.

As with AD, the repurposing approach in PD has gained momentum due to the availability of large-scale health datasets. The Health Data Research UK (HDRUK) initiative, launched during the COVID-19 pandemic, greatly expanded access to primary care data across England, enabling population-scale studies aimed at identifying protective factors and informing public health strategies[17]. Although HDRUK access is still limited to pandemic-related studies on a fee-free basis, the Clinical Practice Research Datalink (CPRD)[18, 19] continues to serve as a powerful platform for epidemiological research. CPRD provides longitudinal, curated observational and prescription data for nearly 19 million alive and currently registered with a general practice that is actively contributing data patients (67 million historically) across the UK. It has already supported investigations into dementia risk in relation to inflammatory biomarkers[20], type 2 diabetes[21], and prescription history[22, 23]. In parallel, resources like the UK Biobank provide complementary diagnostic and genetic data[24], offering an increasingly rich foundation for integrative studies.

In this study, we examined differences in pre-diagnosis observational and prescription data between individuals who went on to develop PD and a matched control cohort, from data curated by CPRD. Specifically, we assembled a cohort of individuals with a PD diagnosis, defining the index date as the date of first recorded diagnosis. Each case was matched with individuals of the same age, sex, and clinical practice with no PD diagnosis at index, forming a retrospective case-control study to explore disease risk factors prior to onset.

Our analysis revealed that certain biometric markers were significantly associated with future PD incidence and helped explain correlations between drug classes and disease onset. We found that these observational data clarified the association of specific drug classes—including the GLP-1 receptor agonists used in type 2 diabetes—with a lower incidence of PD. These findings align with growing evidence suggesting a potential disease-modifying role for GLP1-based therapies in neurodegeneration[25-30].

By combining biometric data with prescription histories, we were able to probe the extent to which observed associations may reflect underlying risk profiles versus the potential protective effects of medications. For example, we observed that biometrics not only mediated the apparent link between drug use and PD but also enhanced the repurposing case for select therapeutics. This dual analysis helps differentiate correlation from possible causation and supports the development of hypotheses around new intervention strategies.

As in the case of AD, our findings underscore the value of integrating electronic health records with rigorous epidemiological study design to identify candidate therapies and better understand the biological landscape preceding PD onset. We hope this work will contribute to the growing evidence base for drug repurposing in PD and inform future clinical and mechanistic studies.

## Methods

### Cohort

This study utilized routinely collected data from the Clinical Practice Research Datalink (CPRD), one of the largest primary care electronic health record databases globally[18, 19]. The CPRD encompasses medical records for approximately 24% of the UK primary care population, providing comprehensive information on prescriptions, clinical diagnoses, laboratory results, and referrals originating from primary care consultations. The dataset is broadly representative of the UK population in terms of size, socio-demographic characteristics, and geographical distribution of practices. Its validity for pharmaco-epidemiological, clinical, and public health research has been well established[20-23].

The study cohort comprised individuals aged 50 years or older who received a first diagnosis of Parkinson’s disease (PD) between January 1, 2012, and December 31, 2022. Each case was individually matched with control patients without PD on age, sex, and primary care general practice, with an average of three controls per case (see Table 1). PD diagnoses were identified using relevant systematized nomenclature of medical clinical terms (SNOMED CT) and Read codes consistent with clinical definitions of the condition. The SNOMED CT Concept/Description Ids are: 49049000/81717011, 49049000/1230677014, 49049000/2920648011, 49049000/1230678016, 49049000/1230679012, 715345007/3302310019. Data were drawn from the CPRD Aurum database version as of September 2023.

**Table 1.** The Parkinson’s disease (PD) case-control (CTL) cohort demographics. Age distributions are given as mean ± standard deviation.

|  | WOMEN |  | MEN |  |
| --- | --- | --- | --- | --- |
| | AGE(mean±-sd) | N | AGE(mean±-sd) | N |
| PD | 75.48±-9.15 | 14284 | 74.62±-9.16 | 23626 |
| CTL | 75.48±-9.15 | 42852 | 74.62±-9.16 | 70878 |

For the observation-based PD propensity prediction, missing data was imputed based on a random selection from data in the same category. Missingness threshold was set at 40% resulting in 9 observations, in addition to age at index date and sex, in which 60% of the data points were complete. A propensity score was generated using the ranger random forest R package (www.rdocumentation.org/packages/ranger). The model was tested based on a 70/30 training/testing data split, and statistics based on multiple runs with diferent splits and imputation. The distribution of observables over the entire PD and control cohort was visualised using a 2-dimensional Uniform Manifold Approximation and Projection (UMAP)[31] using the uwot R package (www.rdocumentation.org/packages/uwot)

## Observations

Observational data were collected for each individual over two retrospective periods: 5 to 10 years and 10 to 20 years prior to the index date, defined as the date of PD diagnosis for cases and the corresponding date for matched controls. For each measure, values were averaged within each of the two periods to reduce the influence of short-term variability. Observations falling outside two standard deviations from the mean were excluded as outliers to improve data quality.

The dataset included 54 routinely recorded observational measures, such as systolic blood pressure (SBP), diastolic blood pressure (DBP), and multiple plasma or serum biomarkers. Not surprisingly, these measures varied in population coverage; for example, SBP was recorded in a substantially larger proportion of individuals compared to laboratory values such as serum albumin.

To maintain the integrity of the matched case–control design, individuals without a complete match on age at PD diagnosis, sex, and primary care general practice were excluded from the relevant biometric analysis.

### Medication

Medication exposure was defined by prescription events for any of the British National Formulary (BNF) (www-medicinescomplete-com.apollo.worc.ac.uk/#/browse/bnf) (1364) medications. All prescription variants of a given BNF type are pooled. Consistent with the observational data, two different time ranges prior to index were analysed, thereby pooling prescription data from 5 to 10 and from 10 to 20 years prior to diagnosis.

### Analysis of Associations

Associations between biometric measures and PD status were quantified using Cohen’s *d* effect sizes. In contrast, associations between drug prescriptions and PD incidence were assessed using odds ratios derived from logistic regression models. Prescription status for each drug was treated as a binary variable, defined by the presence of at least one prescription within the relevant observation period.

For medications predominantly prescribed to one sex—defined as a greater than tenfold difference in prescription frequency between men and women—analyses were restricted to the relevant sex. To adjust for individual differences in overall healthcare engagement and prescribing patterns, the total number of prescriptions was included as a covariate in the regression models.

As a sensitivity analysis, we also incorporated primary care general practice as a random effect in a mixed-effects logistic regression model to account for potential clustering by practice. The inclusion of this random effect and additional covariates did not materially alter the results or conclusions.

To address the possible confounding of biometric risk with prescription type we collected the first prescription date for the given medication classes in the non-PD cohort. We matched individuals with a medication, with those without, based on having the same age at prescription date, sex and primary care general practice. Average levels of the observational measures were the calculated for periods up to five years prior to first prescription. The observation levels prior to prescription in the two groups were compared through a Cohen’s effect size, and a sensitivity analysis with match group randomisation was implemented to test for robustness.

## Results

### Biometric Observations

Analysis of the biometric measures associated with increased risk of developing PD revealed an overall picture very similar to that seen in our previous study for AD (32). The most significant associations are displayed in Figure 1.

**Figure 1.**
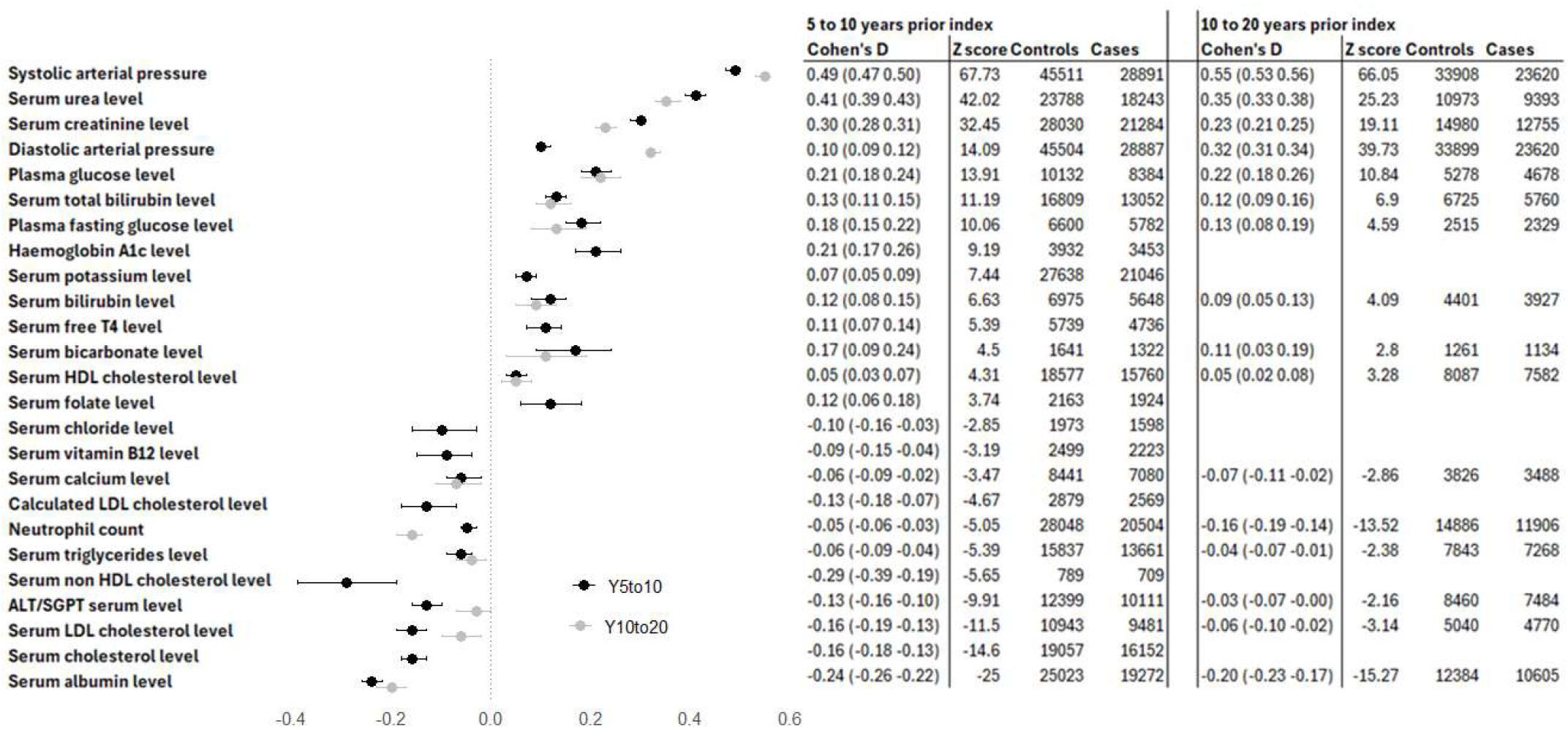
The most significant biometric measure associations with PD for data averaged over 5 to 10 and 10-20 years prior to diagnosis. Data are presented as Cohen’s d effect size (with confidence intervals), where positive effect sizes indicate increased measures, and negative effect sizes reduced measures in PD (Cases) compared to Controls. Z score indicates degree of significance.

These findings point to a complex biomarker profile in the preclinical phase of PD, involving cardiovascular, metabolic, renal, and inflammatory systems. It is of interest then to see to what extent alteration in these easily measurable factors can be combined into a predictive measure to facilitate clinical trial selection or targeted prevention strategies. To this end and as described in the Methods section, we built a random forest predictive model for PD incidence based on these biometric data. The measures had different coverages across the cohort and we built a data matrix from measures with a minimum 60% coverage. With this 40% missingness, we obtained a matrix covering 50,705 individuals across nine observables: Systolic arterial pressure, Diastolic arterial pressure, Serum potassium level, Serum albumin level, Serum creatinine level, Serum urea level, Neutrophil count, Serum sodium level, Serum alkaline phosphatase level. Note, the last two observables in this list did not fall into the top significances for individual measures (Figure 2).

**Figure 2.**
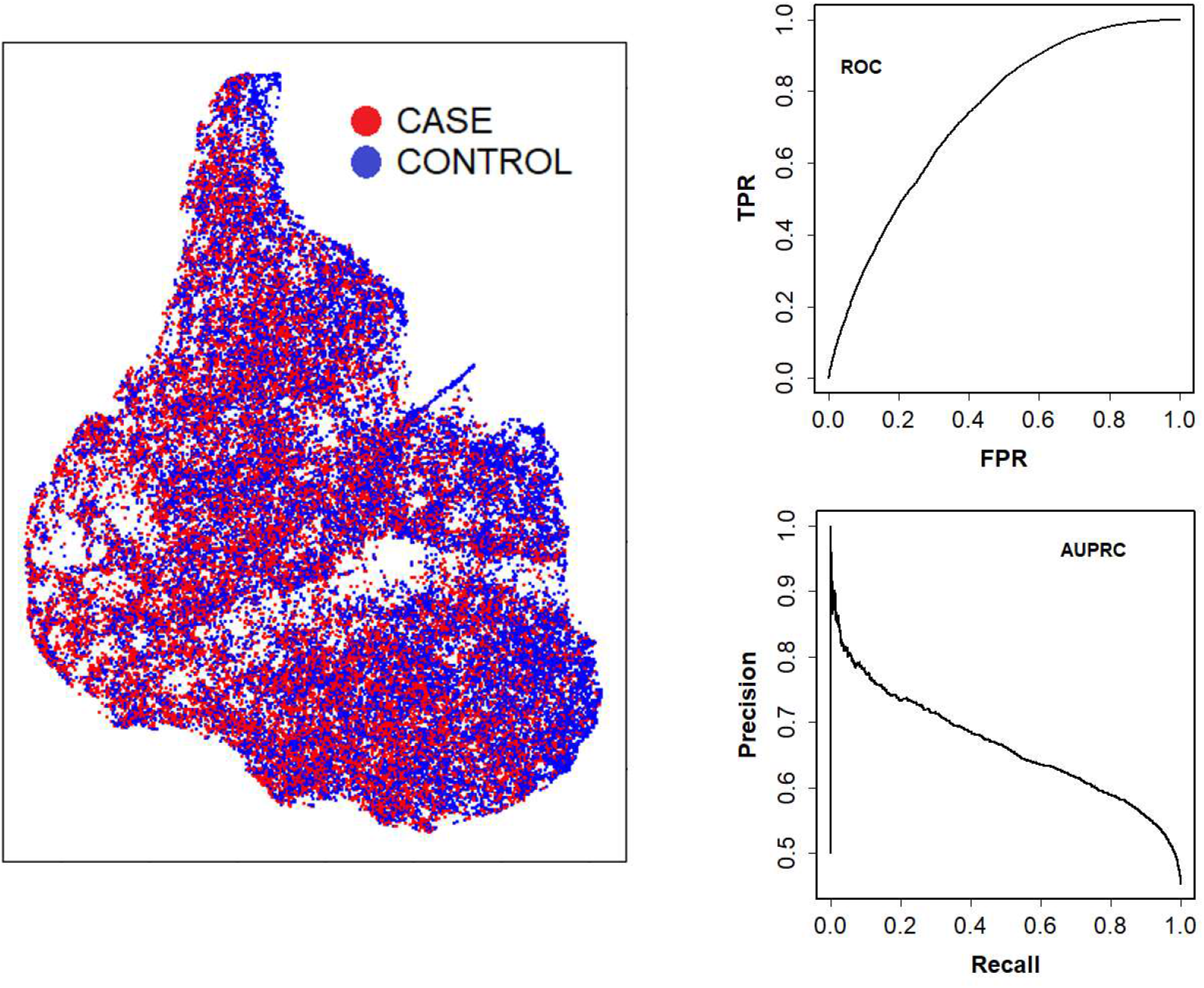
Visualisation and testing of the predictive model. a) A Uniform Manifold Approximation and Projection (UMAP) dimensional reduction plot of biometric variables across the PD case and control cohorts shows the broad overlap in nine key measures at the 5 to 10 years prior index date. A random forest categorisation is moderately effective in segregating the cohorts as illustrated by b) the Receiver Operating Characteristic (ROC) AUC plot of True Positive Rate (TPR) versus False Positive Rate (FPR) and c) the Area Under Precision Recall (AUPR) plot.

The 2-dimensional UMAP plot (Figure 2a), shows broad overlap of the PD (Case) and non-PD (Control) cohorts across these nine key measures at the 5 to 10 year prior index date. It is evident that there is no clear segregation according to future PD diagnosis. However, it is of interest to see to what extent these high coverage observables can be combined into a prediction scheme for PD incidence. To this end we built a random forest model with all nine observables, together with age at index date and sex. The performance of the predictive model is shown as the Receiver Operating Characteristic (ROC) curve for multiple training/testing runs (Figure 1b). The model showed moderate predictability, with the ROC area under curve (AUC) of 0.73±0.003 and an Area Under Precision Recall (AUPR) score of 0.67±0.006. Based on random forest categorical assignments the model has accuracy: 0.67±0.002, sensitivity: 0.64±0.005, and specificity: 0.69±0.004 further indicated moderate predictability of the model. These results are in line with our previous study of AD[32]. By way of comparison, a prediction scheme based on the levels of 20 out of a possible 1536 blood borne proteins, measured over an entire 15-year period prior to PD diagnosis, returned an ROC AUC of 0.8[33]. Our predictive scheme was based on observables with high and some significant measures were thus excluded from the model. We expect that an increase in coverage of the latter measures may result in a more robust predictive model for PD incidence.

### Prescriptions

We next looked at relative drug prescription frequencies prior to index in the case and control cohorts. The results recapitulate the findings of Mittal et al who reported a lower PD incidence in those on the b2 adrenergic receptor agonist salbutamol, based on an analysis of prescription data in the Norwegian population[16]. The results were bolstered by the complementary observation that propranolol, an antagonist of the β2 adrenergic receptor, is associated with a higher PD incidence[16]. We find that both in the 5 to 10 and 10 to 20 years prior to PD diagnosis salbutamol is associated with a lower, and propranolol with a higher, PD incidence (see Table 2). The opposite associations of salbutamol and propranolol noted here and elsewhere point to a target-based effect as salbutamol is an agonist and propranolol an antagonist of the same β2 adrenergic receptor. However, the causal relationship between β2 adrenergic receptor agonism and protection against PD is still a matter of debate. Mechanistically, multiple mechanisms likely contribute. For example, Mittal et al[16] propose a direct effect of β2 agonists on SNCA whereas we [Duty et al[34]] based the proposed repurposing of salbutamol for PD on its driving up the expression of striatal fibroblast growth factor 20 (FGF20).

**Table 2.** PD incidence rates associated with an agonist and antagonist of the b2 adrenergic receptor (salbutamol and propranolol), an alternative corticosteroid asthma medication (beclomethasone) and nicotine.

| DRUG | 5 to 10 years prior PD |  | 10 to 20 years prior PD |  |
| --- | --- | --- | --- | --- |
|  | EFFECT(CI) | Z | EFFECT(CI) | Z |
| SALBUTAMOL | -0.45 (-0.41 -0.49) | -22.1 | -0.53 (-0.49 -0.57) | -24.5 |
| PROPRANOLOL | 0.22 (0.29 0.14) | 5.62 | 0.17 (0.24 0.10) | 4.61 |
| BECLOMETASONE | -0.09 (-0.05 -0.13) | -4.14 | -0.16 (-0.12 -0.20) | -7.55 |
| NICOTINE | -0.85 (-0.75 -0.95) | -16.4 | -0.59 (-0.50 -0.68) | -12.6 |

Subsequent studies have claimed that smoking is a critical confounder of the epidemiological link between asthma medication and PD incidence as smoking is associated with a lower risk of PD and higher likelihood of salbutamol medication[35]. In this context, we also find that nicotine appears to be associated with a lower PD incidence. Interestingly, beclomethasone, an alternative asthma medication with an indirect corticosteroid-based inflammation suppressive activity, also appears to be associated with lower PD incidence (see Table 2). However, the effect size here is much lower than that linking salbutamol with reduced risk, suggesting a potential additional benefit from salbutamol in PD, above and beyond that of its link with asthma. The associations are in line with published data.

Overall, as with the observable biometrics, there is a broad agreement with what we found here in PD, and that previously identified with our AD incidence study[32]. In view of this similarity, for the more extensive analysis we focussed our medication scrutiny on the compound categories we had delimited in the AD study namely drugs for treating hypertension (HT; specifically Angiotensin Converting Enzyme (ACE) inhibitors and Calcium Channel Blockers (CCB)), hormonals, triptans and GLP-1 receptor agonists, see Table 3. Our data revealed that the hypertension medications appear to be associated with significantly higher PD incidence, while the remaining three classes of medication appear to be associated with a significantly reduced incidence of PD (Table 3). The association between hypertension medications and increased PD incidence broadly mirrors that observed in the AD study and points to vascular health being a key factor in both these neurological conditions. GLP-1 receptor agonists are prescribed for T2D which is a high-risk group for PD[36-38]. Our finding of reduced incidence of PD in people taking GLP-1 receptor agonists bolsters the likelihood of these drugs being protective against developing PD. As noted in Figure 2, we also identified plasma glucose elevation as a risk factor for PD, in line with this.

**Table 3.**
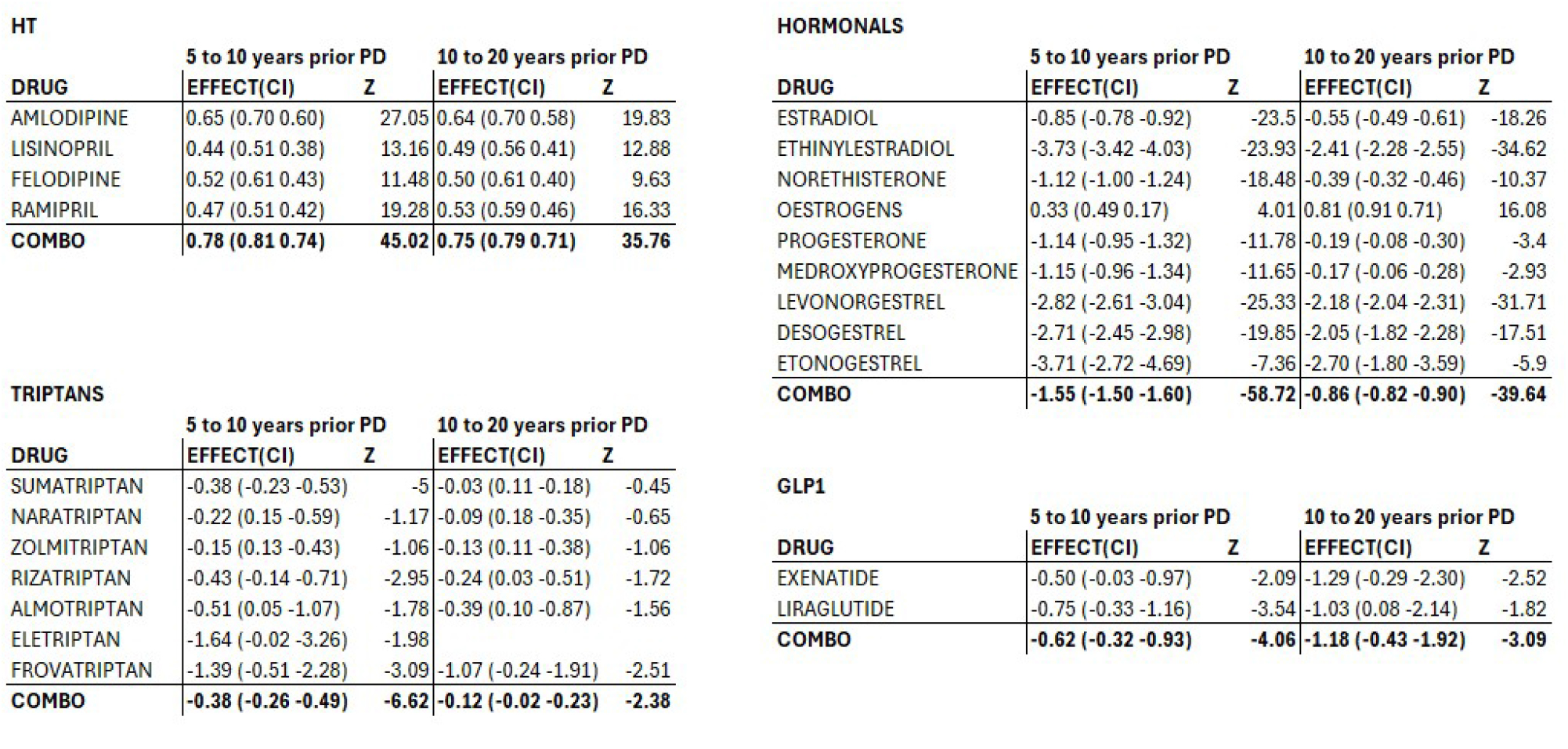
The Associations between PD incidence and the prescriptions of anti-hypertensive medications (HT), hormonal drugs, GLP-1 receptor agonists, and triptans.

These associations with medication alone may, however, be misleading in the absence of an analysis of the underlying PD risk in those being prescribed the drug. To address possible confounding effects of biometric status prior medication we looked at biometric data in the context of future drug prescription within the Control cohort. Specifically, we delimited a prescription group for each medication class with index date corresponding to the earliest prescription date, and a matched control group with no such prescriptions but having the same age, sex and primary care general practice at index. Biometric data was compared between the two cohorts in the five years pre index (Table 4). As a positive control, we observed high systolic blood pressure (SBP) and diastolic blood pressure (DBP) prior to the prescription of anti-hypertensives. This observation explains the higher PD incidence in those on anti-hypertensives. Interestingly, until recently GLP-1 receptor agonists have been prescribed exclusively for type 2 diabetes, which is an established PD risk factor. Our data recapitulates this observation that GLP-1 receptor agonists tend to be prescribed in populations with biometrics associated with higher PD risk (e.g. increased SBP and DBP), see Table 4. The negative association of GLP-1 receptor agonists use with PD incidence is therefore highly significant and our results strengthen the case for repurposing the drug for PD. In contrast, both hormonals and triptans appear to be prescribed in individuals with lower SBP and DBP. Given that low BP is associated with lower PD incidence, these data suggest that the positive associations of both hormonal and triptan medication with reduced PD risk may be explained by cohort differences already apparent prior to prescription, rather than the medications themselves being associated with reduced PD risk.

**Table 4.** Differences in biometric measures between control cohort individuals later prescribed anti-hypertensives (HT), hormonal medication, triptan migraine medication or GLP-1 receptor agonists and matched control cohorts never given these prescriptions. The biometric measures were analysed for the 5-year period prior to index (date of first prescription in control cohort on medication).

| HT |  |  | HORMONALS |  |  |
| --- | --- | --- | --- | --- | --- |
| OBSERVATION | BEFORE MEDICATION |  | OBSERVATION | BEFORE MEDICATION |  |
|  | EFFECT(CI) | Z |  | EFFECT(CI) | Z |
| Systolic arterial pressure | 1.45 (1.41 1.49) | 78.97 | Systolic arterial pressure | -0.92 (-0.99 -0.85) | -28.6 |
| Diastolic arterial pressure | 1.03 (0.99 1.07) | 55.9 | Diastolic arterial pressure | -0.54 (-0.60 -0.47) | -16.5 |
| Serum urea level | 0.26 (0.20 0.32) | 8.69 | Serum urea level | -0.68 (-0.82 -0.54) | -10 |
| Serum creatinine level | 0.20 (0.15 0.25) | 7.71 | Serum creatinine level | -0.45 (-0.57 -0.33) | -7.62 |
| Haemoglobin A1c level | 0.38 (0.27 0.49) | 6.67 | Plasma glucose level | -0.67 (-1.03 -0.31) | -3.78 |
| Serum alkaline phosphatase level | 0.20 (0.14 0.26) | 6.34 | Serum alkaline phosphatase level | -0.18 (-0.32 -0.04) | -2.48 |
| Plasma glucose level | 0.43 (0.27 0.60) | 5.2 | Serum HDL cholesterol level | -0.24 (-0.47 -0.02) | -2.15 |
| Serum triglycerides level | 0.23 (0.14 0.32) | 5.14 | Neutrophil count | 0.13 (0.02 0.23) | 2.32 |
| Plasma fasting glucose level | 0.46 (0.22 0.70) | 3.83 | Serum albumin level | 0.21 (0.08 0.34) | 3.09 |
| Neutrophil count | 0.09 (0.04 0.15) | 3.38 |  |  |  |
| ALT/SGPT serum level | 0.24 (0.10 0.39) | 3.29 | TRIPTANS |  |  |
| Serum potassium level | 0.06 (0.01 0.12) | 2.39 | OBSERVATION | BEFORE MEDICATION |  |
| Serum globulin level | 0.11 (0.00 0.22) | 2.04 |  | EFFECT(CI) | Z |
| HbA1c level | 0.36 (0.01 0.70) | 2.04 | Systolic arterial pressure | -0.49 (-0.55 -0.42) | -14.3 |
| Serum chloride level | -0.26 (-0.47 -0.0) | -2.44 | Serum urea level | -0.66 (-0.78 -0.53) | -11 |
| Serum albumin level | -0.16 (-0.22 -0.1) | -5.37 | Diastolic arterial pressure | -0.22 (-0.29 -0.15) | -6.39 |
|  |  |  | Serum creatinine level | -0.28 (-0.38 -0.17) | -5.33 |
| GLP1 |  |  | Plasma glucose level | -0.74 (-1.10 -0.38) | -4.33 |
| OBSERVATION | BEFORE MEDICATION |  | Haemoglobin A1c level | -0.43 (-0.65 -0.22) | -4.13 |
|  | EFFECT(CI) | Z | Serum alkaline phosphatase level | -0.21 (-0.33 -0.08) | -3.25 |
| Haemoglobin A1c level | 2.35 (1.90 2.80) | 13.13 | Serum globulin level | -0.31 (-0.53 -0.08) | -2.68 |
| Neutrophil count | 0.67 (0.42 0.92) | 5.43 | Serum HDL cholesterol level | -0.19 (-0.37 -0.01) | -2.09 |
| Systolic arterial pressure | 0.48 (0.28 0.69) | 4.97 | Serum albumin level | 0.21 (0.09 0.32) | 3.46 |
| Serum triglycerides level | 0.82 (0.42 1.21) | 4.2 |  |  |  |
| Diastolic arterial pressure | 0.39 (0.19 0.60) | 3.91 |  |  |  |
| Serum alkaline phosphatase level | 0.48 (0.21 0.75) | 3.51 |  |  |  |
| Serum LDL cholesterol level | -0.62 (-1.15 -0.0) | -2.35 |  |  |  |
| Serum cholesterol level | -0.51 (-0.84 -0.1) | -3.03 |  |  |  |
| Serum albumin level | -0.46 (-0.73 -0.1) | -3.38 |  |  |  |
| Serum sodium level | -0.57 (-0.82 -0.3) | -4.64 |  |  |  |
| Serum HDL cholesterol level | -1.24 (-1.59 -0.9) | -7.87 |  |  |  |

## Conclusions

The retrospective analysis of prescription and observational data can inform drug repurposing based on differences in prescription rates in cohorts that will go on to develop the given condition relative to those that are spared. Observational parameters impact on disease susceptibility. Notably, vascular health is a risk factor for developing neurodegenerative disease. Thus, taking account of observational measure differences in prescription rates is critical in building a case for the potential of candidate therapeutics. In this study we gathered prescription and observation data from CPRD for a cohort of 25,000 UK individuals with PD and matched these on age at diagnosis, sex and primary care general practice. Observational differences were evident between the case and control cohorts at 10 to 20 years prior to diagnosis and become more significant at 5 to 10 years prior to diagnosis. The results point to vascular and inflammatory factors driving PD, in agreement with previous reports[39, 40]. These findings are broadly aligned with studies reporting significant biomarker and mechanistic overlap between PD and AD[41-45]. Notably, hypertension is an established risk factor for AD[46-48], serum albumin levels show a negative correlation with brain beta-amyloid depositions[49]. Dysregulation of glucose metabolism[50], and inflammatory pathways[51] have also been reported in AD. It appears that this association analysis can thus be classed as a broad-brush survey of the risk landscape for neurodegenerative conditions as there is very little in the way of disease specific susceptibilities emerging from our basic statistical analysis.

Differences in drug prescription rates between Case and Control cohorts also showed a clear overlap between PD and AD. One striking observation in our analysis of AD was the lower rates of female hormone medication and migraine drug use in the AD cohort. The same picture emerges for our analysis of PD. However, in contrast to the hypertension medication cohort, we find that hormone and migraine prescriptions are higher in those with a lower observational parameter-based PD risk profile. Thus, we argue that these particular drugs may offer no protection against PD and suggest that these data may shed light on the failure of clinical trials of hormonal drugs for other neurodegenerative conditions like dementia[52]. In any case, the possible confounding protective effect of underlying vascular health would have to be carefully controlled for in a future retrospective data selection.

Our observational data does, however, bolster the repurposing potential of GLP-1 receptor agonists for PD. These drugs are historically prescribed for T2D which is a risk factor for both PD and AD. Accordingly, we found that GLP-1 receptor agonists were prescribed at higher rates to control cohorts whose biometric measures, including higher SBP and DBP, were associated with higher PD (and AD) risk. These drugs therefore appear to more than overcome the T2D PD risk, and there is a good case for continuing to explore GLP-1 receptor agonists for PD repurposing, despite recent disappointing outcomes with exenatide in phase III trials[53]. Indeed, positive results have emerged for liraglutide in the context of AD, with the drug sparing grey matter and limiting ventricle growth in phase 2 year long trial of AD patients[30]. The recent licensing of GLP-1 receptor agonists as weight loss drugs[54] may thus positively impact dementia incidence in the future, and our analyses reported here support how they could also do so for PD.

One of the striking observations was the strong negative association of hormonal use in the female population with PD incidence. We questioned whether this reflected a beneficial effect of hormonal use on PD risk, or whether or whether the biometric risk factors we had identified are themselves associated with differences in hormonal prescription likelihood.

When restricting our analysis to the control cohort without PD diagnosis, we found that those on hormonal drugs were more likely to have biometric profiles corresponding to lower PD risk, such as low SBP and DBP. The positive associations of hormonal medication with reduced PD risk are therefore may be explained by cohort differences already apparent prior to prescription, rather than any specific association with PD incidence itself. In this connection, hypertension has been a contraindication for hormone replacement therapy[55-59]. This interpretation would be in line with the Women’s Health Initiative Memory Study (WHIMS) on post 65 year old women that showed no decreased risk of AD in postmenopausal women prescribed hormonals, reviewed by Coker et al[52].

The other medication class associated with a lower PD risk was the triptan migraine medications. Similarly to the hormonal medication cohort, we found that triptans tend to be prescribed to those with lower biometric risk of developing PD, implying the drugs themselves have may have no protective effects against PD. However, there have been reports of protective effects of zolmitriptan in a rat model of PD[60] and sumatriptan in a mouse stroke model[61], so further scrutiny is required here.

One possible future iteration of retrospective data analysis emerging from this work is the potential to construct virtual clinical trials from observational health record data, incorporating the hypothesised causal inference frameworks. By first using biometric profiles to stratify individuals into risk groups for PD, we can define a high-risk cohort that mimics the eligibility criteria of a traditional trial. Within this cohort, those prescribed a given drug (e.g., GLP-1 receptor agonists) can be compared to matched individuals not prescribed the drug, controlling for confounders such as age, sex, comorbidities, and baseline biometric markers. This emulated trial design, akin to a target trial, allows estimation of the comparative effectiveness of medications in delaying or reducing PD incidence. The approach offers a pragmatic pathway to prioritise therapeutic candidates before prospective trials and may be particularly valuable in diseases with long prodromal phases like PD. Applying this framework to the present dataset, especially for promising candidates like GLP-1 receptor agonists, could strengthen the evidence base for disease-modifying potential and refine stratification for future interventional studies.

### Study limitations

As with most retrospective studies, it is problematic to make conclusions as to causality based on correlations. For example, as shown here, the negative association with PD incidence of hormonal drugs and anti-migraine medication may be confounded by their higher prescription rates in those with biometric profiles falling in the lower PD risk group. In contrast to the analysis of biometric factors, where one is comparing numerical measures, the analysis of medication incidence is complicated by being categorical, where a null assignment for a given medication may be due to the lack of data, rather than a person being medication naive. We have mitigated against this by controlling for overall prescription frequency but ideally would balance the case and control groups on general overall medication load.

## Data Availability

The data is available from the CPRD under appropriate licence agreement

## Acknowledgements

The author would like to thank Dr Alexadru Dregan for extracting the data from CPRD and helping with technical issues. This study is based in part on data from the Clinical Practice Research Datalink obtained under licence from the UK Medicines and Healthcare products Regulatory Agency. The data is provided by patients and collected by the NHS as part of their care and support. The interpretation and conclusions contained in this study are those of the authors alone.

## Ethical approval

The CPRD group obtained ethical approval from a National Research Ethics Service Committee for all purely observational research using anonymised CPRD data, as in the present study. The study protocol has been approved by the Independent Scientific Advisory Committee for Medicines and Healthcare Products Regulatory Agency, Ref: 14_111.

## Abbreviations

PD: Parkinson’s Disease
AD: Alzheimer’s Disease
SBP: Sistolic Blood Pressure
DBP: Diastolic Blood Pressure
CPRD: Clinical Practice Research Datalink
HDRUK: Health Data Research UK
SNOMED CT: systematized nomenclature of medical clinical terms
UMAP: Uniform Manifold Approximation and Projection
BNF: British National Formulary
ROC: Receiver Operating Characteristic
AUC: Area Under Curve
AUPR: Area Under Precision Recall
TPR: True Positive Rate
FPR: False Positive Rate
SNCA: alpha-synuclein
FGF20: fibroblast growth factor 20
ACE: Angiotensin Converting Enzyme
CCB: Calcium Channel Blockers
T2D: Type 2 Diabetes
WHIMS: Women’s Health Initiative Memory Study

